# COVID-19 impact on consecutive neurological patients admitted to the emergency department

**DOI:** 10.1101/2020.05.23.20110650

**Authors:** Andrea Pilotto, Alberto Benussi, Ilenia Libri, Stefano Masciocchi, Loris Poli, Enrico Premi, Antonella Alberici, Enrico Baldelli, Sonia Bonacina, Laura Brambilla, Matteo Benini, Salvatore Caratozzolo, Matteo Cortinovis, Angelo Costa, Stefano Cotti Piccinelli, Elisabetta Cottini, Viviana Cristillo, Ilenia Delrio, Massimiliano Filosto, Massimo Gamba, Stefano Gazzina, Nicola Gilberti, Stefano Gipponi, Marcello Giunta, Alberto Imarisio, Paolo Liberini, Martina Locatelli, Francesca Schiano di Cola, Renata Rao, Barbara Risi, Luca Rozzini, Andrea Scalvini, Veronica Vergani, Irene Volonghi, Nicola Zoppi, Barbara Borroni, Mauro Magoni, Matilde Leonardi, Gianluigi Zanusso, Sergio Ferrari, Sara Mariotto, Alessandro Pezzini, Roberto Gasparotti, Ciro Paolillo, Alessandro Padovani

**Affiliations:** Department of Clinical and Experimental Sciences, Neurology Unit, University of Brescia, Italy; Parkinson’s disease Rehabilitation unit, FERB Onlus Trescore Balneario, Bergamo, Italy; Stroke Unit, ASST Spedali Civili di Brescia, Italy; Department of Neuroimmunology and Neuromuscular Diseases, Foundation IRCCS Neurological Institute Carlo Besta, Milan, Italy; Neurology Unit, School of Medicine, University of Bologna, Bologna, Italy; Neurophysiology Unit, ASST Spedali Civili, Brescia, Italy; Neurology, Public Health, Disability Unit – Fondazione IRCCS Istituto Neurologico Carlo Besta, Milan, Italy; Department of Neurosciences, Biomedicine and Movement Sciences, Neurology Unit, University of Verona, Verona; Neuroradiology Unit, University of Brescia and ASST Spedali Civili of Brescia, Brescia, Italy; Emergency Department ASST Spedali Civili di Brescia, Brescia, Italy

**Keywords:** COVID-19, emergency department, neurology, stroke, encephalitis

## Abstract

**Objective:** Aim of this study was to analyse the impact of COVID-19 on clinical and laboratory findings and outcome of neurological patients consecutively admitted to the emergency department (ED) of a tertiary hub center.

**Methods:** All adult patients consecutively admitted to the ED for neurological manifestations from February 20^th^ through April 30^th^ 2020 at Spedali Civili of Brescia entered the study. Demographic, clinical, and laboratory data were extracted from medical records and compared between patients with and without COVID-19.

**Results:** Out of 505 consecutively patients evaluated at ED with neurological symptoms, 147 (29.1%) tested positive for SARS-CoV-2. These patients displayed at triage higher values of CRP, AST, ALT, and fibrinogen but not lymphopenia (p<0.05). They were older (73.1 ± 12.4 vs 65.1 ± 18.9 years, p=0.001) had higher frequency of stroke (34.7% vs 29.3%), encephalitis/meningitis (9.5% vs 1.9%) and delirium (16.3% vs 5.0%). Compared to patients without COVID, they were more frequently hospitalized (91.2% vs 69.3%, p<0.0001) and showed higher mortality rates (29.7% vs 1.8%, p<0.0.001) and discharge disability, independently from age.

**Conclusions:** COVID-19 impacts on clinical presentation of neurological disorders, with higher frequency of stroke, encephalitis and delirium, and was strongly associated with increased hospitalisation, mortality and disability.

## Introduction

The outbreak of coronavirus disease 2019 (COVID-19), caused by the severe acute respiratory syndrome coronavirus 2 (SARS-CoV-2), hit Italy by the end of February and rapidly spread from Lombardy to the rest of the country, with a number of fatalities beyond 31,000. Although all regions have reported having patients with COVID-19, the highest number of identified cases was in the provinces of eastern Lombardy[1]. With the increasing number of confirmed cases and the accumulating clinical data, it is now well established that, in addition to the predominant respiratory symptoms, a significant proportion of COVID-19 patients has central and peripheral neurological symptoms, including olfactory dysfunction[2], headache and confusion[3]. Several case reports and small series also suggested an association between COVID-19 and cerebrovascular events[4], immune-mediated peripheral[5] and central nervous system involvement[6-8].

To date, however, no comprehensive large surveys on the impact of COVID19 on neurological status of patients presenting at the emergency room has been published[4, 6,9]. In fact, still little is known on which are the most common acute neurological presentations in COVID-19 and whether they differ compared to patients without COVID-19 in terms of severity and outcomes. This knowledge is pivotal for the organisation of neurological services and for better use of triage limited resources during the COVID-19 pandemic.

Indeed, in our experience, the urgent drift to cope with the rapidly overwhelming number of simultaneously critical patients drew the conversion of the majority of neurological units into non-specialistic wards for broader and generic COVID-19 patient care. The converted units became spokes referring neurological patients to tertiary hub centers for specialty care. This led to the definition in tertiary centers of dedicated medical and nurse teams working in isolated spaces (Special Neuro-COVID Units) in order to guarantee equal access to acute therapies.

The aim of this study is to investigate the impact of COVID-19 on patients with neurological diseases by recording clinical presentations, laboratory characteristics and management/outcomes of a series of neurological patients who consecutively presented at the emergency department (ED) during the peak of the epidemic. The study was carried out in a tertiary referral neurological center, the first Italian Neuro-COVID unit, identified as a hub for stroke and neurological emergencies in an area of more than 1,200,000 people that, to date, is hosting the greatest number of admitted COVID-19 neurologic patients in Italy[1].

## Methods

### Study design and participants

The observational study included all adult patients evaluated for neurological symptoms at the Emergency Department (ED) of the Hospital “Azienda Socio Sanitaria Territoriale (ASST) Spedali Civili”, Brescia, Italy. The study was approved by the local ethics committee of ASST Spedali Civili di Brescia Hospital (NP 4067, approved 08.05.2020).

### Data collection and definitions

The medical form and admission data of included patients ranged from February 20^th^ through April 30^th^ 2020. Epidemiological, demographical, clinical, laboratory and outcome data were extracted from medical records using standardised anonymised data collection forms. All data were imputed and checked by five physicians (AP, AB, IL, SM, MG).

SARS-CoV-2 detection in respiratory specimens were performed by real-time RT PCR methods. Both nasopharyngeal and oropharyngeal swabs were performed in all patients. If two consecutive tests obtained at least 24 hours apart resulted negative, and there was high suspicion of COVID-19 (i.e. interstitial pneumonia at chest x-ray, low arterial partial pressure of oxygen), a bronchoalveolar lavage was performed.

Standardized blood examinations at ED included complete blood count, serum biochemical tests including C-reactive protein and liver profile.

### Statistical analysis

Continuous and categorical variables are reported as mean with standard deviation and n (%) respectively. Differences between patients with and without COVID-19 were compared by t-tests, χ^2^ test or Fisher’s exact test and multivariate model adjusted for age and sex where appropriate. A two-sided *p*<0.05 was considered statistically significant. Data analyses were carried out using SPSS software (version 22.0)

## Results

Five hundred and five adult patients with neurologic symptoms were evaluated at the ED starting from February 20th through April 30 2020. Of these, 147 (29.1%) were found positive for SARS-CoV-2. Demographical, clinical and laboratory characteristics of patients are reported in **Table 1**.

**Table 1.**
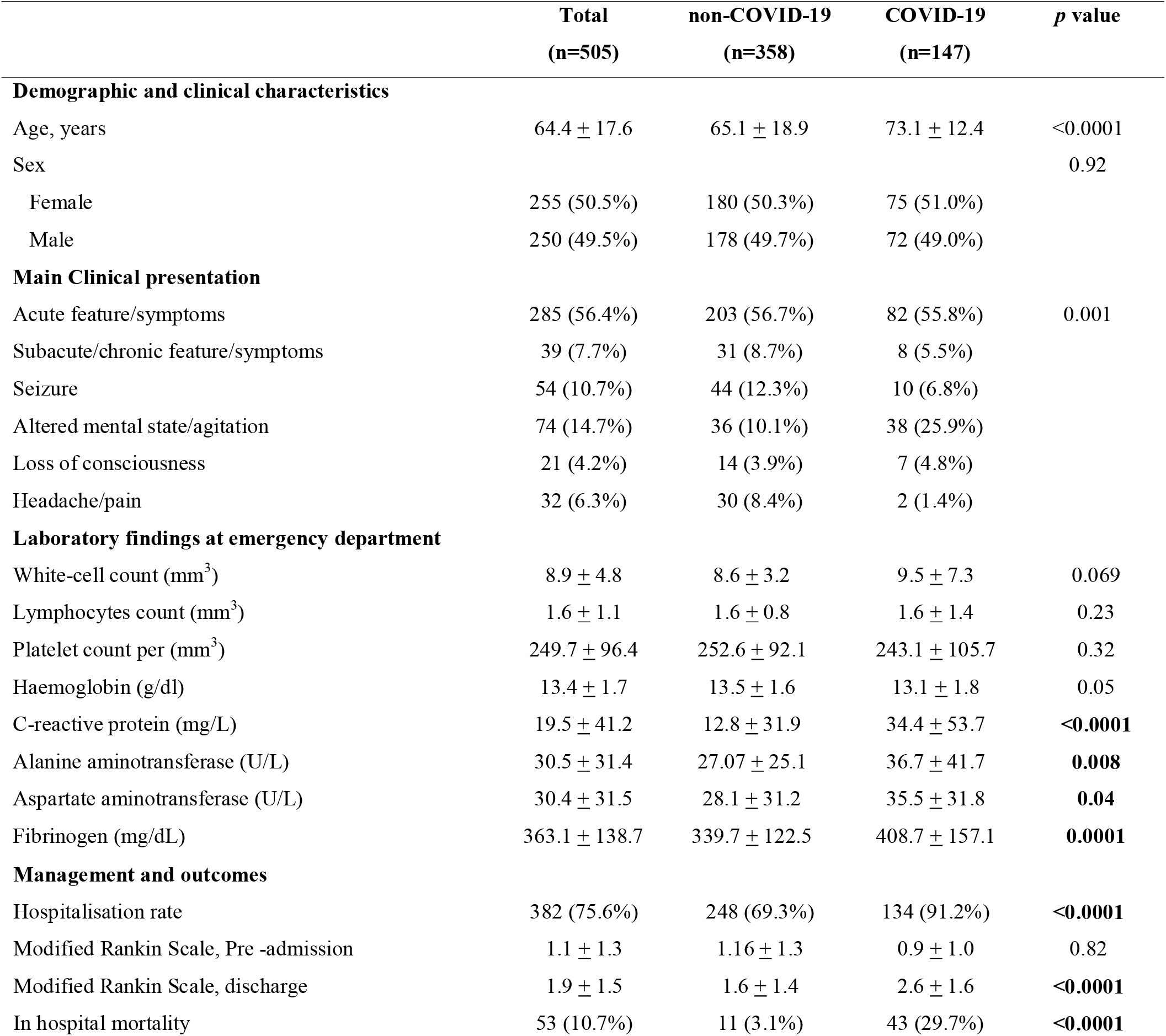
Demographic, clinical, laboratory characteristics and clinical management of neurological patients evaluated at the emergency department. Data are mean, or n (%). p values were calculated by T-test or Fisher’s exact test, as appropriate.

|  | Total<br>(n=505) | non-COVID-19<br>(n=358) | COVID-19<br>(n=147) | p value |
| --- | --- | --- | --- | --- |
| **Demographic and clinical characteristics** |  |  |  |  |
| Age, years | 64.4 ± 17.6 | 65.1 ± 18.9 | 73.1 ± 12.4 | <0.0001 |
| Sex |  |  |  | 0.92 |
| Female | 255 (50.5%) | 180 (50.3%) | 75 (51.0%) |  |
| Male | 250 (49.5%) | 178 (49.7%) | 72 (49.0%) |  |
| **Main Clinical presentation** |  |  |  |  |
| Acute feature/symptoms | 285 (56.4%) | 203 (56.7%) | 82 (55.8%) | 0.001 |
| Subacute/chronic feature/symptoms | 39 (7.7%) | 31 (8.7%) | 8 (5.5%) |  |
| Seizure | 54 (10.7%) | 44 (12.3%) | 10 (6.8%) |  |
| Altered mental state/agitation | 74 (14.7%) | 36 (10.1%) | 38 (25.9%) |  |
| Loss of consciousness | 21 (4.2%) | 14 (3.9%) | 7 (4.8%) |  |
| Headache/pain | 32 (6.3%) | 30 (8.4%) | 2 (1.4%) |  |
| **Laboratory findings at emergency department** |  |  |  |  |
| White-cell count (mm<sup>3</sup>) | 8.9 ± 4.8 | 8.6 ± 3.2 | 9.5 ± 7.3 | 0.069 |
| Lymphocytes count (mm<sup>3</sup>) | 1.6 ± 1.1 | 1.6 ± 0.8 | 1.6 ± 1.4 | 0.23 |
| Platelet count per (mm<sup>3</sup>) | 249.7 ± 96.4 | 252.6 ± 92.1 | 243.1 ± 105.7 | 0.32 |
| Haemoglobin (g/dl) | 13.4 ± 1.7 | 13.5 ± 1.6 | 13.1 ± 1.8 | 0.05 |
| C-reactive protein (mg/L) | 19.5 ± 41.2 | 12.8 ± 31.9 | 34.4 ± 53.7 | <0.0001 |
| Alanine aminotransferase (U/L) | 30.5 ± 31.4 | 27.07 ± 25.1 | 36.7 ± 41.7 | 0.008 |
| Aspartate aminotransferase (U/L) | 30.4 ± 31.5 | 28.1 ± 31.2 | 35.5 ± 31.8 | 0.04 |
| Fibrinogen (mg/dL) | 363.1 ± 138.7 | 339.7 ± 122.5 | 408.7 ± 157.1 | 0.0001 |
| Management and outcomes |  |  |  |  |
| Hospitalisation rate | 382 (75.6%) | 248 (69.3%) | 134 (91.2%) | <0.0001 |
| Modified Rankin Scale, Pre -admission | 1.1 ± 1.3 | 1.16 ± 1.3 | 0.9 ± 1.0 | 0.82 |
| Modified Rankin Scale, discharge | 1.9 ± 1.5 | 1.6 ± 1.4 | 2.6 ± 1.6 | <0.0001 |
| In hospital mortality | 53 (10.7%) | 11 (3.1%) | 43 (29.7%) | <0.0001 |

Compared to COVID negative patients, COVID+ were significantly older (73.1 ± 12.4 vs 65.1 ± 18.9 years, p=0.001) and had a different distribution of neurological presentation (p=0.001), with a higher frequency of altered mental status or delirium (25.9% vs 10.1%) but lower frequency of seizures and headache/pain (see Table 1 for details). According to final diagnosis, COVID+ had higher prevalence of ischaemic stroke (n=51, 34.7% vs n=105, 29.3%), delirium (n=24, 16.3% vs n=18, 5.0%) and meningitis/encephalitis (n=14, 9.5% vs n=7, 1.9%) (Supplementary Table 1, Figure 1).

**Figure 1.**
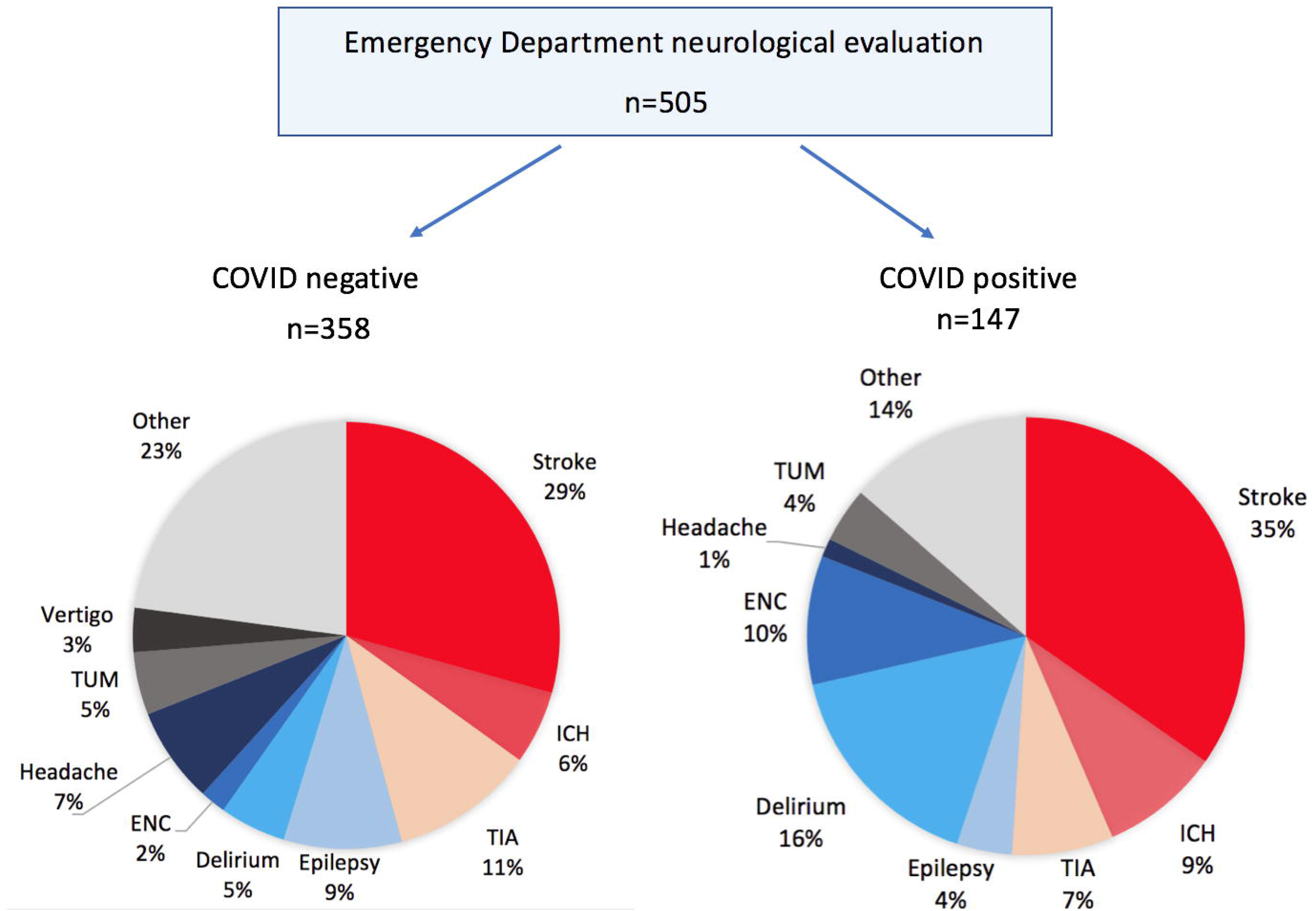
Different diagnostic distribution in neurological patients with COVID and without COVID at the Emergency Department. Abbreviations: COVID, COVID-19 disease; ENC, encephalitis/encephalitis; ICH, intracranial Haemorrhage, TIA, transient ischemic attack; TUM, Central Nervous system tumour or metastasis

Laboratory analysis in COVID+ patients showed increased serological levels of C-reactive protein, aspartate and alanine aminotransferase and fibrinogen (all p<0.05) but no differences were observed for whole white blood cell count, lymphocytes, haemoglobin, or platelet count (all p>0.05, see Table 1).

COVID+ showed a higher rate of hospitalisation following ED triage (91.2% vs 69.3%, p<0.0001) and in-hospital mortality (29.7% vs 3.1%, p<0.0001). In particular, mortality was higher in ICH (COVID+ 58.3% vs nonCOVID 33.0%), in ischemic stroke (COVID+ 39,2% vs nonCOVID 2,8%) and in delirium/altered mental status (COVID+23,4% vs nonCOVID 0%).

According to modified Rankin scale, COVID+ patients had higher levels of disability at discharge compared to nonCOVID patients (2.6 ± 1.6 vs 1.6 ± 1.4, p<0.0001) though both groups had similar baseline values (Table 1).

## Discussion

This study evaluated the impact of SARS-CoV-2 infections on clinical manifestation in patients with neurological symptoms consecutively admitted to the ED during the COVID-19 outbreak in northern Italy. Findings showed that about a third of neurological patients assessed at the ED resulted positive for SARS-CoV-2 and that these were characterised by an increased frequency of cerebrovascular events and encephalitis (Figure 1). The higher frequency of cerebrovascular events is in line with early observations in China[3,4] and with the known pro-inflammatory state associated with increased risk of both ischaemic stroke and ICH[9]. In fact, in this series, COVID-19 neurological patients showed significantly higher levels of C-reactive protein and fibrinogen, strongly arguing for endothelial dysfunction or cardioembolic events as possible risk factors for cerebrovascular accidents, as reported by recent works[10,11].

Diagnosis of encephalitis were also more frequently observed in COVID-19 patients. Given the close time relationship with SARS-coV-2 infection, a direct or immune-mediated involvement of COVID-19 of the Central Nervous System (CNS) has been hypothesised in agreement with several case descriptions[7,11]. The CNS-potential diffusivity of COVID-19 by blood or nasal-pharyngeal route, as suggested by in vitro-models and described for similar viral agents, is still debated[4] although clinical observations of frequent and persistent anosmia/dysgeusia in COVID-19 infected subjects[2] has been claimed as supportive evidence.

In this series, COVID-19 neurological patients had a unfavourable outcome as shown by increased rates of hospitalisation, high rate of mortality and higher disability at discharge. The mortality rate observed is far higher compared to general in-hospital mortality reported in COVID-19 both in China[10,13] and in Western countries[14] and is higher compared to non COVID-19 cases. This further highlight the severe impact of COVID-19 on neurological patients, being these patients more fragile, vulnerable and thus particularly prone to early deterioration and death.

This study, evaluating more than five hundred patients admitted at the ED, extended previous reports[4,6,15] and highlighted the fact that COVID-19 infection specifically affects clinical presentations and management of neurological patients. This has deep implications for daily practice and healthcare organization. For neurological patients severe enough to warrant hospitalization, our experience suggests that the creation of Neuro-COVID units might represent the most suitable solution able to provide urgent, standardized and tailored neurologic care.

We acknowledge that this study entails some limitations. First, due to the study design we limited the observation to patients presenting with prominent neurological manifestations thus probably underestimating neurological symptoms such as headache/pain or confusion in more severe patients, as suggested in previous reports[2,4,6]. Second, interpretation of findings could be limited by the single-centre design and by the re-distribution of emergency network in the region, potentially leading to decrease access to healthcare systems for less severe neurological manifestations.

In conclusion, this study has shown that one third of neurological patients admitted to emergency department were SARS-CoV-2 positive and that most of them were affected by cerebrovascular disorders and altered mental status due to encephalitis or severe delirium. Coronavirus plays a relevant impact on health status as neurological patients required more frequently hospitalization, and were at higher risk of mortality and disability at discharge. Further studies are needed to compare different models of care in order to identify the most efficacious organisation to counteract the impact of COVID-19 on neurological patients.

## Data Availability

Data are made available from authors upon reasonable request

## Author contributions

Conception and design of the study: AP, AB and AP. Acquisition and analysis of data: AP, AB, IL, SM, LP, EP, AP, EB, SB, LB, MB, SC, MC, AC, SCP, EC, VC, ID, MF, MG, SG, NG, SG, MG, AI, PL, ML, FS, RR, BR, LR, AS, VV, IV, NZ, BB, MM, mL, GZ, SF, SM, AP, RB, CP and AP. Drafting the manuscript and figures: AP, AB and AP.

## Financial disclosure and conflict of interest regarding the research related to the manuscript

All authors have no conflict of interest regarding the research related to the manuscript. The full financial disclosure for the past year (for all authors) is documented at the end of the manuscript.

